# Impact of the first COVID-19 shelter-in-place order in the United States on emergency department utilization, Marin County, California

**DOI:** 10.1101/2020.07.01.20144691

**Authors:** Brett R. Bayles, Michaela F. George, Haylea Hannah, Patti Culross, Rochelle R. Ereman, Dustin W. Ballard, Matthew Willis

## Abstract

**Background:** The first shelter-in-place (SIP) order in the United States was issued across six counties in the San Francisco Bay Area to reduce the impact of COVID-19 on critical care resources. We sought to assess the impact of this large-scale intervention on emergency departments (ED) in Marin County, California.

**Methods:** We conducted a retrospective descriptive and trend analysis of all ED visits in Marin County, California from January 1, 2018 to May 4, 2020 to quantify the temporal dynamics of ED utilization before and after the March 17, 2020 SIP order.

**Results:** The average number of ED visits per day decreased by 52.3% following the SIP order compared to corresponding time periods in 2018 and 2019. Both respiratory and non-respiratory visits declined, but this negative trend was most pronounced for non-respiratory admissions.

**Conclusions:** The first SIP order to be issued in the United States in response to COVID-19 was associated with a significant reduction in ED utilization in Marin County.

## INTRODUCTION

The World Health Organization (WHO) designated an outbreak of a novel coronavirus (COVID-19) as a global public health emergency on January 30, 2020.^1^ By early March, the first COVID-19 associated deaths in the United States were identified in California.^2^ As healthcare systems braced for an influx in demand for critical care resources due to life threatening respiratory symptoms,^3^ communities began implementing non-pharmaceutical interventions such as physical distancing to slow the spread of the disease. The first-in-the-country shelter-in-place (SIP) order was issued across six counties in the San Francisco Bay Area on March 17, 2020 and became a state-wide mandate in California on March 19.^4^

Mitigation efforts designed to limit physical contact in the population have likely had an early impact on disease transmission patterns,^5^ but empirical evidence is needed to assess the specific effects of these interventions on healthcare systems.^6^ While the number of COVID-19 cases and hospitalizations increased in many locations during the early pandemic period,^7^ decreases in emergency medical services for other serious health conditions have been observed.^8-11^ Quantifying the changing trends in healthcare utilization is critical for assessing the public health impact of SIP orders and other COVID-19 mitigation efforts, particularly given the possibility of future epidemic waves.^12^

To better understand the impact of early COVID-19 mitigation efforts on medical systems, we measured trends in emergency department (ED) utilization before and after the first-in-the-country SIP order was issued in Marin County, California.

## METHODS

Marin County has the highest percentage of residents over age 65 among the six Bay Area counties to first issue SIP orders,^13^ and may be at increased risk for higher rates of COVID-19 hospitalizations and fatalities.^14^ We obtained surveillance data on daily ED visits across three acute care facilities from the Marin County Department of Health and Human Services. We then conducted a retrospective temporal trend analysis of all ED visits reported in the county from January 1, 2018 to May 4, 2020.

Demand for emergency medical services may differ according to type of health condition.^10-11^ Therefore, we dichotomized ED visits as either respiratory or non-respiratory based on syndromes derived from chief complaints (i.e. primary symptoms a patient states as their reason for seeking medical care in the ED). Criteria was based on initial COVID-19 symptoms reported by the Centers for Disease Control and Prevention in March 2020 and expert clinical knowledge from the Marin County Public Health Officer and Emergency Medical Services Director (Table S1).

We analyzed the average number of daily ED visits between January 1 through May 4 in 2018, 2019, and 2020 to establish a comparative baseline and assess the impact of SIP order on ED utilization. We compared a series of discrete time periods during the early pandemic period (January 1-May 4) in 2020 with the same intervals in 2018 and 2019. The percent change in total, respiratory, and non-respiratory ED visits was then calculated between 2020 and the combined average of the corresponding baseline time periods in 2018 and 2019. We used the Mann-Kendall non-parametric test for monotonic trends to detect changes in the magnitude and direction (i.e. consistently increasing or decreasing) daily ED visits within each time period. Statistical analysis was conducted using R version 3.2.3.^15^

## RESULTS

Marin County is a primarily suburban/rural county with a relatively stable population of 260,000, which is supported by three acute care facilities averaging 63,781 ED visits per year. Our results show a substantial decrease in ED visits associated with the first-in-the-country SIP order issued on March 17, 2020. Compared to the same time intervals in 2018 and 2019, the average number of daily ED visits in 2020 decreased 17.6% between March 3-16, and 52.3% between March 17-31 [mean (SD) = 148.8 (17.7) and 86.7 (13.8), respectively] (Table). The decrease in ED volume relative to the previous baseline time periods persisted between April 1-14 [mean (SD) = 78.8 (7.1)] and April 15-May 4 [mean (SD) = 97.6 (13.1)].

**Table 1.** Differences in daily ED visits between 2018 through 2020 in Marin County, California.

|  | Time Periods |  |  |  |  |
| --- | --- | --- | --- | --- | --- |
|  | Jan 1-Mar 2 | Mar 3-16 | Mar 17-31 | April 1-14 | April 15-May 4 |
| Total ED visits, avg. (SD) |  |  |  |  |  |
| 2018 | 146.0 (42.9) | 174.3 (12.9) | 173.5 (28.4) | 181.0 (15.0) | 178.1 (15.7) |
| 2019 | 175.7 (16.7) | 186.6 (16.7) | 190.0 (18.5) | 148.1 (33.2) | 184.2 (28.1) |
| 2020 | 166.8 (13.4) | 148.8 (17.7) | 86.7 (13.8) | 78.8 (7.1) | 97.6 (13.1) |
| Change, % | +3.7 | -17.6 | -52.3 | -52.1 | -46.1 |
| Respiratory ED visits, avg. (SD) |  |  |  |  |  |
| 2018 | 11.1 (6.2) | 8.2 (3.2) | 6.8 (2.9) | 7.5 (2.4) | 5.6 (2.5) |
| 2019 | 8.4 (3.2) | 10.5 (3.0) | 10.4 (4.2) | 7.29 (2.3) | 7.2 (2.1) |
| 2020 | 10.6 (3.5) | 11.43 (2.38) | 7.33 (3.77) | 4.71 (2.27) | 3.4 (1.7) |
| Change, % | +8.6 | +22.2 | -14.8 | -36.3 | -46.9 |
| Non-respiratory ED visits, avg. (SD) |  |  |  |  |  |
| 2018 | 134.9 (39.9) | 166.1 (11.7) | 166.7 (27.4) | 173.5 (15.1) | 172.5 (15.0) |
| 2019 | 167.3 (15.56) | 176.1 (16.7) | 179.6 (15.8) | 140.79 (32.44) | 177.0 (26.9) |
| 2020 | 156.2 (13.6) | 137.36 (17.4) | 79.3 (11.5) | 74.1 (7.0) | 94.2 (13.1) |
| Change, % | +3.4 | -19.7 | -54.28 | -52.9 | -46.1 |

The SIP order was associated with a decline in both non-respiratory and respiratory ED visits, but the change in average ED volume was more substantial for non-respiratory visits (Figure 1). Between March 17-31, respiratory visits decreased by 14.8% and non-respiratory visits decreased by 54.2% compared to the same time periods in the previous two years.

**Figure 1.**
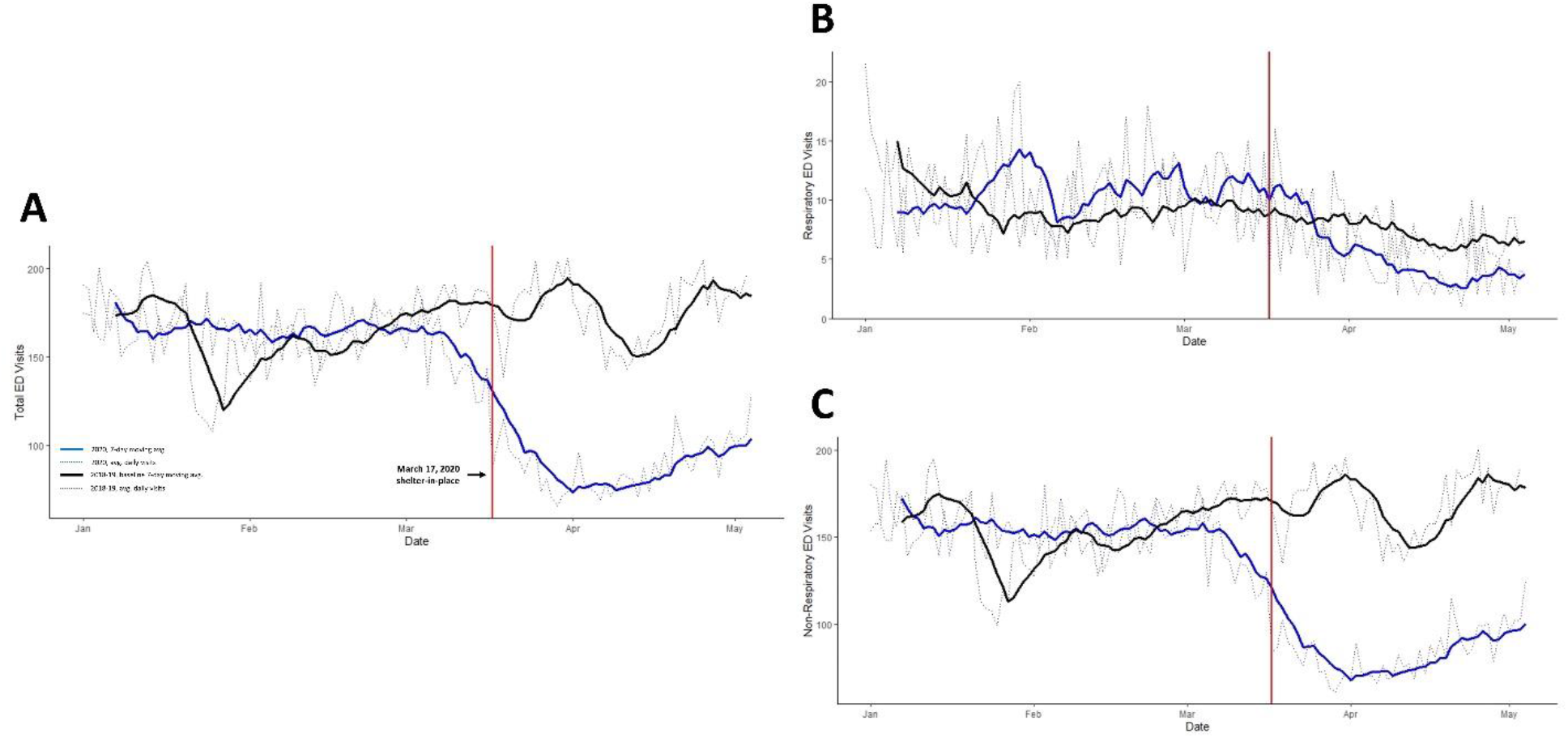
Temporal trends in emergency department utilization in 2020 (blue line) compared to previous two-year average 2018-19 baseline (black line) for A) all emergency department visits, B) respiratory emergency department visits, and C) non-respiratory emergency department visits. Vertical red line denotes March 17 shelter-in-place order in Marin County.

We found statistically significant temporal trends in daily ED visits within several of the early pandemic time intervals (Table S2). Between March 3-16 and March 17-31, total daily ED visits decreased at a statistically significant rate (p<0.05). Again, the magnitude of the negative trend was greater for non-respiratory visits compared to respiratory visits after the SIP order.

## DISCUSSION

We found evidence that the first-in-the-country SIP order was associated with a significant reduction in ED visits in Marin County. Compared to the same baseline time periods over the last two years, the decrease in volume of ED visits was largest for non-respiratory symptoms. These trends are consistent with other observations, which suggests those with serious medical conditions not related to COVID-19 are not seeking medical care.^8-11^

This study has several strengths. These analyses were able to utilize previous data from 2018 and 2019 as a comparison for change, the data is inclusive of all EDs in Marin County, and over 140,000 visits were included. However, the limitations include the inability to verify symptoms based on clinical chart review, due to limited data available from ED records. Because the data came from three different EDs, the coding of chief complaints may have varied.

Diverging trends in healthcare utilization have emerged during the early COVID-19 pandemic period. Our results suggest that SIP orders and other circumstances of the COVID-19 pandemic were associated with reduced ED utilization. This reduction may be due to public fear or other unintended consequences of reduced human mobility (e.g. fewer traffic accidents), and reduced transmission of other communicable diseases due to reduced physical contact. Future directions include a time series analysis to better understand the statistical impacts of SIP orders in Marin, further stratification of syndromes to separate causes of non-respiratory visits, and investigation of linking ED visit data with other data sources such as emergency medical services database, death records, and the human mobility index.

In conclusion, COVID-19 mitigation efforts including large-scale SIP orders can significantly impact healthcare systems. Specifically, ED utilization appears to be closely associated with these orders and appear very responsive to temporal trends in disease dynamics. This information may be used to support clinical-decision making and guide strategic public health messages if future non-pharmacological interventions are required.

## Data Availability

Data may be made available at the discretion of the corresponding author.

## Author Contributions

All authors contributed to the concept and design of study, drafting and critical revision of the manuscript. BB conducted the statistical analysis and revised the final draft.

## Ethics approval

This retrospective temporal analysis of ED data was reviewed by the University of Washington ethics review board, who determined that these activities do not meet the regulatory definition of research as the Marin County Department of Health and Human Services collaborated with these researchers for assistance in analyzing and interpreting ED data to help understand the impacts of the COVID-19 pandemic and associated policies in Marin County and to protect the public’s health.

